# Obsessive-Compulsive Symptoms and Its Association with Psychological Strain Among Chinese University Students

**DOI:** 10.1101/2025.08.01.25332558

**Authors:** Ziyan Chen, Jiaqi Yang, Xiner Li, Yongsheng Li, Yuwen Huang, Harold G. Koenig, Zhizhong Wang, Lunchi Huang

## Abstract

**Objectives:** Obsessive-compulsive symptoms (OCS) are commonly seen in non-clinical populations but receive considerably less attention compared to obsessive-compulsive disorder and other mental health issues. This study examined the prevalence and characteristics of OCS and its relationship with psychological strain among Chinese college students.

**Methods:** A cross-sectional online survey was conducted in April 2024 involving 3,322 participants; the Yale-Brown Obsessive-Compulsive Scale and Coping Strain scale were administered.

**Results:** The prevalence of OCS was 6.5%, non-medical major students (OR, 1.471; 95%CI, 1.027-2.017) were at a higher prevalence risk than medical students. Participants with stronger OCS were statistically and positively associated with the higher severity of coping strain (OR, 5.115; 95%CI, 3.438-7.609; p < 0.001).

**Conclusion:** OCS and psychological strain may mutually contribute to the maintenance and/or reinforcement of each other among Chinese college students, suggesting a new interventional model that may help to promote mental health among this population.

## Introduction

Obsessive-compulsive disorder (OCD) is a mental health disorder characterized by the presence of persistent obsessions and/or compulsions that affect 1% to 3% of the global population (Brock et al., 2025) and 1.6% of adults in China (Huang et al., 2019). OCD can cause significant disability, with the World Health Organization (WHO) ranking it as one of the 10 most debilitating conditions (Bobes et al., 2001). Meanwhile, young adults represent a significant proportion of new OCD diagnoses (Sharma et al., 2021), with an increasing trend (from 1.9% in 2009 to 2.4% in 2015) among college students (Oswalt et al., 2020).

It is reported that obsessive-compulsive symptoms (OCS) are seen in 80% of non-clinical populations. These do not amount to a disorder by themselves (Rachman and de Silva, 1978), and the average duration of untreated illness has been estimated to be 17 years (Hollander et al., 1998). A recent study indicated that OCD patients’ symptoms are not fundamentally distinct from those in normal populations, and OCS receives very limited awareness by society (Hu et al., 2023). In China, a study using the SCL-90 showed that OCS scores were significantly higher in 2015 compared to those collected in 1986 (Liu et al., 2018). A cross-national survey found that OCD onset averages at 17.9 years, typically emerging in late adolescence and aligning with university entry in China (Brakoulias et al., 2017). One study reported that the prevalence of OCS measured by the Obsessive-Compulsive Inventory-Revised was 24.1% among Chinese college students (Liu et al., 2020). Meanwhile, the emergence of the COVID-19 pandemic and the subsequent containment measures had profound impacts on individual and societal daily lives (Chaturvedi et al., 2021). There is a lack of studies focusing on the OCS during and post-pandemic among university students.

In the Strain Theory of Suicide and Mental Disorder, a psychological strain must consist of at least two conflicting facts, and if the two facts are non-contradictory, there would be no strain (Zhang et al., 2014). The motivation to control obsessive-compulsive symptoms and the inability to achieve such control constitute two contradictory forces, which generate psychological strain. Individuals with OCS often experience increased risk of suicidal thoughts and behaviors among university students (Hu et al., 2023), although how it increases the risk of suicidal behaviors is not well known (Huz et al., 2016). One possible explanation can be understood through the strain theory. Psychological strain may cause greater distress in individuals with OCS and affect the nature of these symptoms. More research is needed on the relationship between OCS and psychological strain, particularly in non-clinical samples.

This study aims to a) estimate the prevalence of OCS among university students in China; b) describe the characteristics of OCS in Chinese university students; and c) explore the association between obsessive-compulsive symptoms and psychological strain.

## Methods

### Participants and procedure

Participants (undergraduates, postgraduates, and doctoral candidates) were recruited through Chinese social media, email lists, and electronic newsletters. Inclusion criteria were: 1) those currently enrolled in an undergraduate program or higher in mainland China; 2) no major physical disease; and 3) voluntary participation in the study. Exclusion criteria were: 1) non-full-time enrolled students; 2) those with a clinically diagnosed severe psychotic illness; and 3) those incapable of independently using electronic devices to complete online questionnaires due to visual impairment or other disabilities.

Four universities from each of China’s four major regions (East, Central, West, and Northeast) were selected based on their willingness to participate. Students were provided a link to an online questionnaire through WeChat (one of China’s most popular social media platforms). Those who responded to the invitation were encouraged to forward the invitation letter to peers and post it on social media sites. The invitation letter was initially sent to 6,213 potential participants by the WeChat network, of whom 4,341 responded to the invitation; 30 participants refused after reading the informed consent form, resulting in 4,311 completed questionnaires (Fig. 1). Of those, 989 records were excluded during the data cleaning process, leaving a final sample of 3322 participants in the analysis.

**Figure 1.**
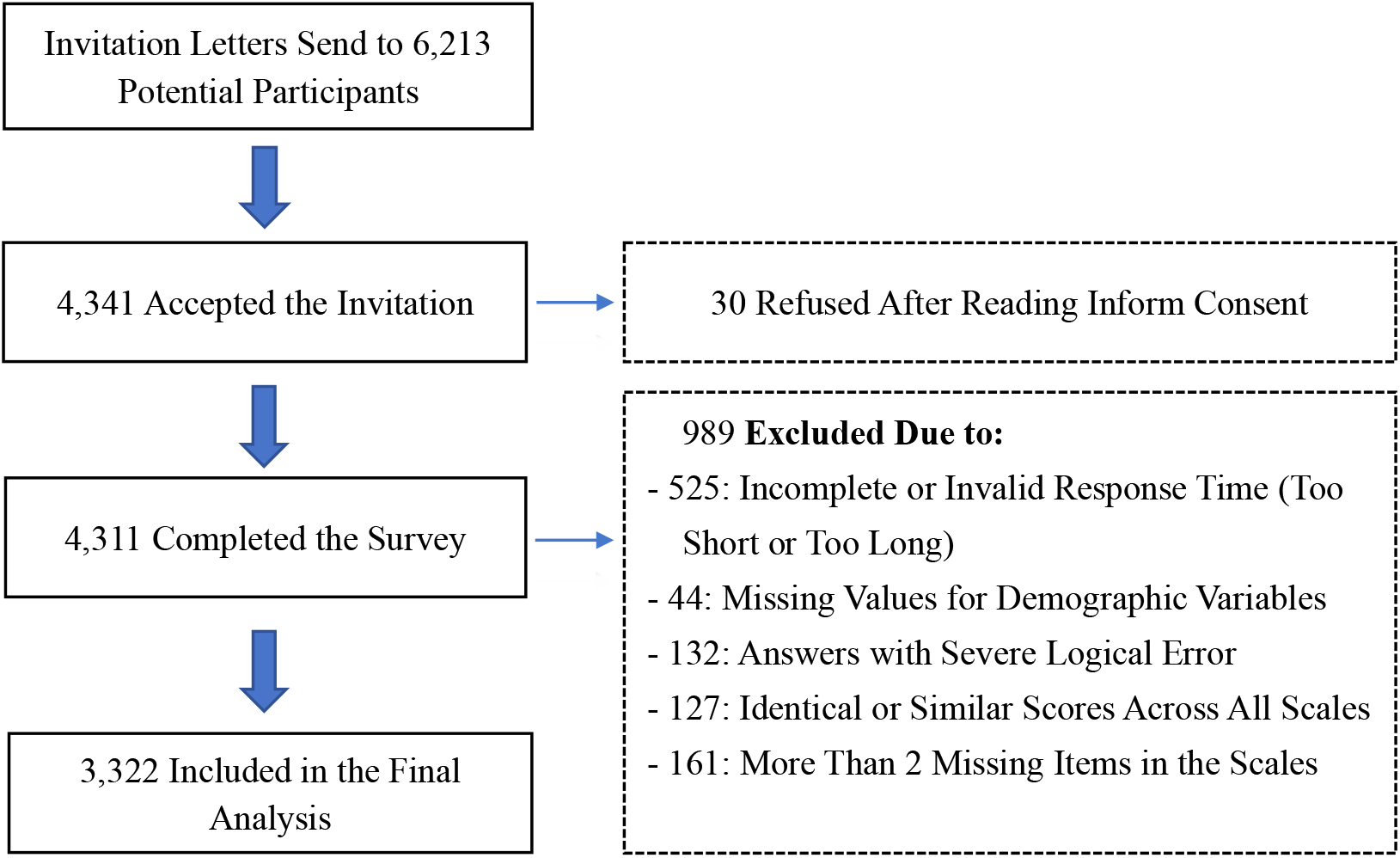
The flowchart of the participants’ enrollment.

The authors assert that all procedures contributing to this work comply with the ethical standards of the relevant national and institutional committees on human experimentation and with the Helsinki Declaration of 1975, as revised in 2008. The survey was anonymous, and online informed consent was obtained by asking participants to check a box on the device’s screen with the response (I agree to participate in the study, or I do not agree to participate in the survey). A small amount of money was offered for their time when the questionnaire finished. The study was approved by the institutional review board of Ningxia Medical University (approval #2017-171, 2020-112).

### Measures

#### Demographic characteristics

Information was collected on age, gender, living area, living city line, ethnicity (Han versus minorities), grade, major, marital status, number of siblings, self-scaled academic performance (poor, moderate, or good), self-scaled family economic status (poor, moderate, or good), hours of sleep, late sleeping behavior (before versus after 23:00) and family history of OCD.

#### Obsessive-compulsive symptom severity

The Yale-Brown Obsessive-Compulsive Scale (Y-BOCS) was originally developed by Goodman et al (1989), and is a clinician-administered instrument designed to evaluate the severity of obsessive-compulsive symptoms. The instrument has five items for obsessions and five items for compulsions, each item is rated on a 5-point Likert Scale ranging from 0 (no symptoms) to 4 (extreme symptoms), with the total score ranging from 0 to 40 correspondingly. Higher scores indicate greater severity of obsessive-compulsive symptoms; ≥ 16 was considered OCS positive. The Chinese version has high internal consistency and fair 1-week test-retest reliability in Chinese-speaking samples (Zhang et al., 2019).

#### Coping strain

The psychological strain was assessed using the 40-item Psychological Strain Scale (PSS), which includes four dimensions: value strain, aspiration strain, deprivation strain, and coping strain. Therefore, only this dimension was included in this study to save time and improve the quality of questionnaire responses. The score of each item ranges from 1 to 5 (1 = strongly disagree, 2 = disagree, 3 = maybe, not sure, 4 = agree, and 5 = strongly agree). Higher scores indicate higher psychological strain (X. Zhang et al., 2017). Previous studies have shown that the PSS has good reliability and validity among Chinese college students, PSS ≥ 25 indicate positive (Zhang et al., 2020).

### Statistical Analyses

Descriptive statistics were performed by calculating means with standard deviations (SD) for Y-BOCS scores and frequencies with percentages for categorical variables. Frequency distributions were compared across three symptom subgroups: obsessive-only, compulsive-only, and combined obsessive-compulsive. The chi-square test was used to test the difference in the prevalence of OCS (Y-BOCS ≥ 16) among demographic variables. Binary logistic regression models were used to test the association after controlling for covariates. The adjusted ORs and their 95% confidence intervals (CIs) were calculated. The analyses were conducted using IBM SPSS version 23.0 software (SPSS Inc., Chicago, IL, USA). All statistical tests were two-tailed, with a type I error rate set at 0.05.

## Results

### Sociodemographic characteristics of the participants

As presented in Table 1. The sample comprised 3,322 Chinese university students, with 16.7% first-year, 23.3% second-year, 26.8% third-year, 13.0% fourth-year, 11.1% fifth-year undergraduates, and 9.1% postgraduates. A majority of participants were female (67.7%), from medical university (69.4%), of Han ethnicity (92.8%), and unmarried (99.5%). About a quarter (23.6%) of students had no siblings. 28.4% perceived their family economic status as good, 62.1% considered as moderate, and 9.5% as poor.

**Table 1.** The demographic characteristics and psychological strain of university students by Y-BOCS score (N = 3322)

| **Variables** | **Total (%)** | **Y-BOCS (Mean ± SD)** | **Y-BOCS $\geq 16$ (n = 215) (%)** | **$\chi^2$** | ***p*** |
| --- | --- | --- | --- | --- | --- |
| Age (year) |  |  |  | 38.419 | < 0.001 |
| $\leq 19$ | 709 (21.3) | 5.47 ± 5.619 | 44 (6.2) | | |
| 20 | 691 (20.8) | 6.02 ± 6.056 | 52 (7.5) | | |
| 21 | 694 (20.9) | 5.80 ± 6.127 | 55 (7.9) | | |
| 22 | 484 (14.6) | 4.88 ± 5.629 | 25 (5.2) | | |
| $\geq 23$ | 744 (22.4) | 4.48 ± 5.694 | 39 (5.2) | | |
| Gender |  |  |  | 13.331 | 0.004 |
| Male | 1072 (32.3) | 4.77 ± 5.726 | 55 (5.1) | | |
| Female | 2250 (67.7) | 5.62 ± 5.910 | 160 (7.1) | | |
| Living city line |  |  |  | 23.189 | 0.006 |
| First-tier city | 620 (18.7) | 5.86 ± 6.294 | 60 (9.7) | | |
| New first-tier city | 912 (27.5) | 5.14 ± 5.738 | 55 (6.0) | | |
| Second-tier city | 618 (18.6) | 5.02 ± 5.510 | 25 (4.0) | | |
| Third-tier city | 1172 (35.3) | 5.41 ± 5.894 | 75 (6.4) | | |
| Ethnicity |  |  |  | 6.137 | 0.105 |
| Han | 3083 (92.8) | 5.27 ± 5.858 | 95 (6.3) | | |
| Minorities | 239 (7.2) | 6.31 ± 5.875 | 20 (8.4) | | |
| Grade |  |  |  | 56.670 | < 0.001 |
| 1 Year | 554 (16.7) | 5.50 ± 5.774 | 37 (5.5) | | |
| 2 Year | 774 (23.3) | 5.93 ± 5.925 | 52 (6.7) | | |
| 3 Year | 890 (26.8) | 5.72 ± 5.884 | 65 (7.3) | | |
| 4 Year | 431 (13.0) | 5.20 ± 6.240 | 32 (7.4) | | |
| 5 Year | 370 (11.1) | 3.51 ± 5.044 | 14 (3.8) | | |
| Postgraduate | 303 (9.1) | 4.94 ± 5.757 | 15 (5.0) | | |
| Major |  |  |  | 25.433 | < 0.001 |
| Medicine | 2306 (69.4) | 5.06 ± 5.629 | 122 (5.3) | | |
| Others | 1016 (30.6) | 6.00 ± 6.320 | 93 (9.2) | | |
| Marital status |  |  |  | 2.179 | 0.536 |
| Unmarried | 3305 (99.5) | 5.36 ± 5.861 | 214 (6.5) | | |
| Married | 17 (0.5) | 3.29 ± 6.233 | 1 (5.9) | | |
| Only child or not |  |  |  | 9.429 | 0.024 |
| No | 2538 (76.4) | 5.44 ± 5.821 | 160 (6.3) | | |
| Yes | 784 (23.6) | 5.03 ± 5.997 | 55 (7.0) | | |
| Academic performance |  |  |  | 7.743 | 0.258 |
| Poor | 188 (5.7) | 5.46 ± 6.394 | 18 (9.6) |  |  |
| Moderate | 2403 (72.3) | 5.33 ± 5.750 | 144 (6.0) |  |  |
| Good | 731 (22.0) | 5.37 ± 6.098 | 53 (7.3) |  |  |
| Living expenses |  |  |  | 21.342 | 0.002 |
| More than needed | 943 (28.4) | 5.22 ± 5.908 | 59 (6.3) |  |  |
| Enough | 2064 (62.1) | 5.18 ± 5.754 | 124 (6.0) |  |  |
| Inadequate | 315 (9.5) | 7.79 ± 6.254 | 32 (10.2) |  |  |
| Hours of sleep |  |  |  | 35.178 | < 0.001 |
| Less than 6 hours | 277 (8.3) | 6.52 ± 6.933 | 39 (14.1) |  |  |
| 6~7 hours | 1827 (55.0) | 5.45 ± 5.813 | 113 (6.2) |  |  |
| 7~8 hours | 1028 (30.9) | 4.92 ± 5.552 | 49 (4.8) |  |  |
| More than 8 hours | 190 (5.7) | 4.92 ± 6.069 | 14 (7.4) |  |  |
| Stay up late or not |  |  |  | 2.503 | 0.475 |
| No | 207 (6.2) | 4.88 ± 5.683 | 9 (4.3) |  |  |
| Yes | 3115 (93.8) | 5.38 ± 5.876 | 206 (6.6) |  |  |
| Family history of OCD |  |  |  | 200.204 | < 0.001 |
| No | 2527 (76.1) | 4.51 ± 5.392 | 113 (4.5) |  |  |
| Yes | 37 (1.1) | 13.32 ± 5.180 | 12 (32.4) |  |  |
| Don't know | 758 (22.8) | 7.73 ± 6.436 | 90 (11.9) |  |  |

### The prevalence of OCS

The probable prevalence of OCS was 6.5% (215/3322). Females had higher Y-BOCS scores and a higher prevalence of OCS. Those who studied non-medical majors, resided in first-tier cities, were from only-child families, with poor academic performance, with inadequate living expenses, and slept less than 6 hours, and the 21-year-old group had a higher prevalence of OCS. Besides, those with a family history of OCD had a higher prevalence of OCS (*p* < 0.001).

### Distribution of the different OCS Symptoms

As shown in Table 2, the probable prevalences for obsessive-only, compulsive-only, and combined obsessive-compulsive symptoms were 4.27% (142/3322), 2.92% (97/3322), and 5.15% (171/3322), respectively. Female students had higher prevalences across all symptom categories. Those sleeping less than 6 hours had a remarkably higher prevalence of mixed obsessive-compulsive symptoms than those sleeping longer than 6 hours. Students with a family history of OCD also had a higher prevalence in all three categories.

**Table 2.**
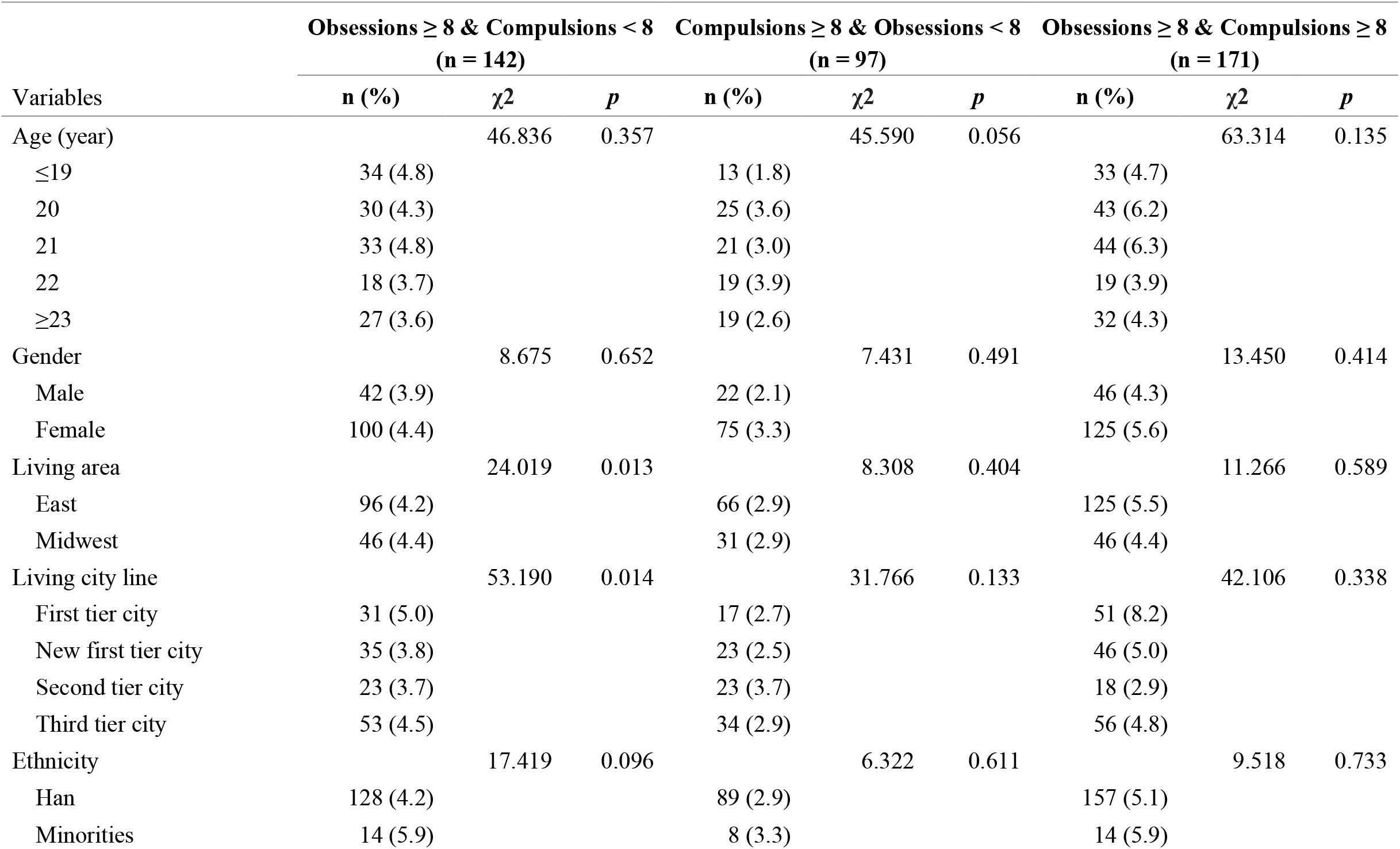
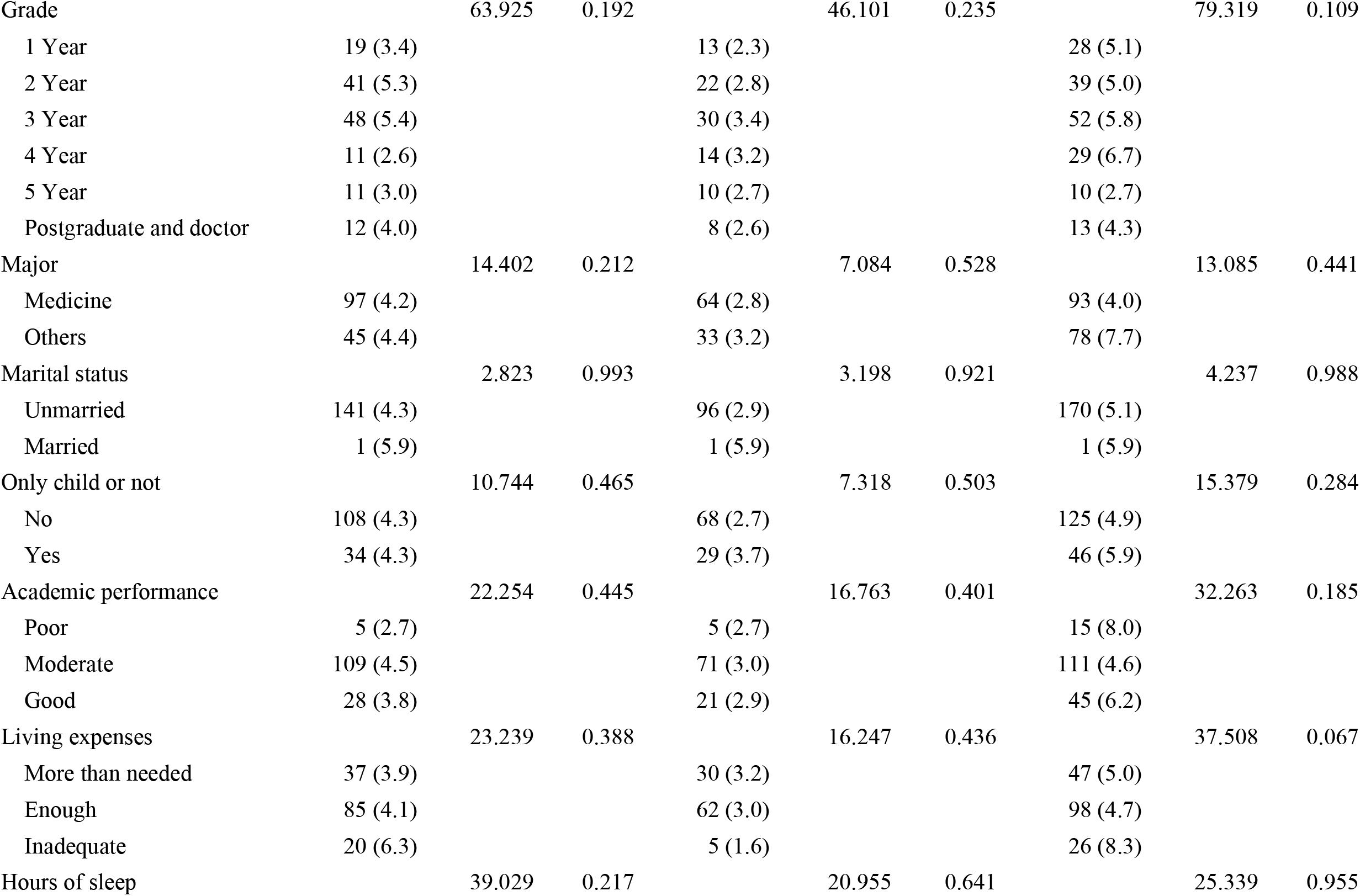
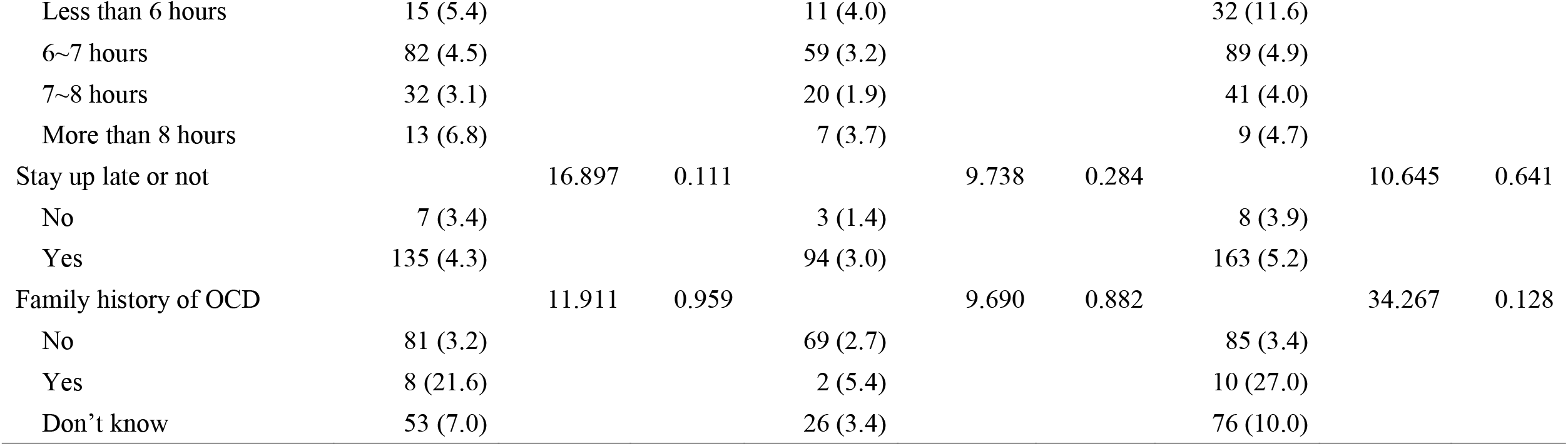
Frequency distribution of demographic and behavioral characteristics across different obsessive-compulsive symptom categories.

### Logistic Regression Analysis

A logistic regression model of obsessive-compulsive symptoms (OCS) included the variables presented in Table 3. Results showed that non-medical major students (OR, 1.471; 95%CI, 1.027-2.017) were at higher prevalence risk than their demographic counterparts. The family history of OCD was significantly associated with OCS (OR, 9.784; 95%CI, 4.446-21.533; *p* < 0.001). Compared with those living in the first-tier city, students living in second-tier (OR=0.440; 95%CI, 0.260-0.745) and new first-tier cities(OR=0.600; 95%CI, 0.394-0.914) have a lower prevalence risk of OCS.

**Table 3.** Logistic regression analysis of factors associated with OCS (N=3322)

| Variables | $\beta$ | SE | Wald $\chi^2$ | *p* | OR (95%CI) |
| --- | --- | --- | --- | --- | --- |
| Age Group ($\leq 19$ ref) | | | 2.275 | 0.685 | |
| 19-20 | 0.320 | 0.278 | 1.323 | 0.250 | 1.377 (0.798-2.373) |
| 20-21 | 0.385 | 0.326 | 1.395 | 0.238 | 1.470 (0.776-2.784) |
| 21-22 | 0.143 | 0.400 | 0.129 | 0.720 | 1.154 (0.527-2.525) |
| $\geq 23$ | 0.341 | 0.446 | 0.585 | 0.444 | 1.406 (0.587-3.368) |
| Gender (female) | 0.266 | 0.176 | 2.285 | 0.131 | 1.305 (0.924-1.843) |
| Living area (east) | 0.100 | 0.192 | 0.272 | 0.602 | 1.105 (0.759-1.609) |
| Living city line (first tier line ref) |  |  | 11.137 | 0.011 |  |
| New first-tier city | -0.511 | 0.215 | 5.651 | 0.017 | 0.600 (0.394-0.914) |
| Second-tier city | -0.820 | 0.268 | 9.330 | 0.002 | 0.440 (0.260-0.745) |
| Third-tier city | -0.303 | 0.218 | 1.944 | 0.163 | 0.738 (0.482-1.131) |
| Ethnicity (Han) | -0.426 | 0.268 | 2.528 | 0.112 | 0.653 (0.386-1.104) |
| Grade (Postgraduate ref) |  |  | 3.785 | 0.581 |  |
| 1 Year | 0.630 | 0.532 | 1.403 | 0.236 | 1.878 (0.662-5.328) |
| 2 Year | 0.235 | 0.476 | 0.242 | 0.623 | 1.264 (0.497-3.217) |
| 3 Year | 0.258 | 0.433 | 0.355 | 0.551 | 1.294 (0.554-3.023) |
| 4 Year | 0.477 | 0.395 | 1.458 | 0.227 | 1.612 (0.743-3.498) |
| 5 Year | 0.114 | 0.406 | 0.079 | 0.779 | 1.121 (0.506-2.482) |
| Major (non-medicine) | 0.386 | 0.161 | 5.731 | 0.017 | 1.471 (1.027-2.017) |
| Marital status (unmarried) | -0.649 | 1.096 | 0.350 | 0.554 | 0.523 (0.061-4.484) |
| Only child or not (yes) | 0.079 | 0.176 | 0.203 | 0.653 | 1.083 (0.766-1.530) |
| Academic performance (moderate ref) |  |  | 2.251 | 0.325 |  |
| Poor | 0.281 | 0.285 | 0.969 | 0.325 | 1.324 (0.757-2.315) |
| Good | 0.228 | 0.181 | 1.578 | 0.209 | 1.256 (0.880-1.791) |
| Living expenses (enough ref) |  |  | 2.863 | 0.239 |  |
| More than needed | 0.071 | 0.178 | 0.161 | 0.688 | 1.074 (0.758-1.522) |
| Inadequate | 0.377 | 0.223 | 2.862 | 0.091 | 1.458 (0.942-2.257) |
| Hours of sleep (less than 6 hours ref) |  |  | 10.325 | 0.016 |  |
| 6~7 hours | -0.620 | 0.212 | 8.551 | 0.003 | 0.538 (0.355-0.815) |
| 7~8 hours | -0.719 | 0.243 | 8.746 | 0.003 | 0.485 (0.303-0.785) |
| More than 8 hours | -0.500 | 0.351 | 2.020 | 0.155 | 0.607 (0.305-1.208) |
| Stay up late or not (yes) | 0.335 | 0.374 | 0.801 | 0.371 | 1.398 (0.671-2.909) |
| Family history of OCD (no ref) |  |  | 49.503 | < 0.001 |  |
| Yes | 2.281 | 0.402 | 32.116 | < 0.001 | 9.784 (4.446-21.533) |
| Don't know | 0.781 | 0.155 | 25.221 | < 0.001 | 2.183 (1.610-2.961) |

Besides, as shown in Table 4, females (OR, 1.413; 95%CI, 1.206-1.655; *p* < 0.001) had a higher risk of coping strain than males. Non-medical major students (OR, 1.217; 95%CI, 1.031-1.436), those living in eastern China (OR, 1.240; 95%CI, 1.039-1.479) were at higher risk compared to their demographic counterparts. Regarding academic performance, students with poor (OR, 2.343; 95%CI, 1.641-3.347; *p* < 0.001) or moderate (OR, 1.430; 95%CI, 1.196-1.711; *p* < 0.001) academic performance had higher risks of OCS than those with good performance. Participants with stronger OCS (OR, 5.115; 95%CI, 3.438-7.609; *p* < 0.001) were statistically and positively associated with the higher severity of psychological strain.

**Table 4.** Logistic regression analysis of factors associated with psychological strain (N=3322)

| Variables | $\beta$ | SE | Wald $\chi^2$ | *p* | OR (95%CI) |
| --- | --- | --- | --- | --- | --- |
| Age Group ($\geq 23$ ref) | | | 13.396 | 0.009 | |
| $\leq 19$ | 0.184 | 0.215 | 0.730 | 0.393 | 1.202 (0.789-1.831) |
| 19-20 | 0.078 | 0.190 | 0.171 | 0.679 | 1.082 (0.746-1.568) |
| 20-21 | 0.092 | 0.173 | 0.282 | 0.596 | 1.096 (0.781-1.538) |
| 21-22 | -0.348 | 0.150 | 5.353 | 0.021 | 0.706 (0.526-0.948) |
| Gender (female) | 0.346 | 0.081 | 18.373 | < 0.001 | 1.413 (1.206-1.655) |
| Living area (east) | 0.215 | 0.090 | 5.712 | 0.017 | 1.240 (1.039-1.479) |
| Living city line (first tier line ref) |  |  | 2.569 | 0.463 |  |
| New first-tier city | 0.135 | 0.114 | 1.393 | 0.238 | 1.144 (0.915-1.430) |
| Second-tier city | 0.193 | 0.126 | 2.339 | 0.126 | 1.213 (0.947-1.553) |
| Third-tier city | 0.100 | 0.117 | 0.734 | 0.392 | 1.105 (0.879-1.388) |
| Ethnicity (Han) | 0.188 | 0.146 | 1.658 | 0.198 | 1.207 (0.907-1.606) |
| Grade (1 year ref) |  |  | 5.046 | 0.410 |  |
| 2 Year | 0.000 | 0.140 | 0.000 | 0.999 | 1.000 (0.761-1.315) |
| 3 Year | 0.138 | 0.168 | 0.676 | 0.411 | 1.148 (0.826-1.594) |
| 4 Year | 0.167 | 0.206 | 0.654 | 0.419 | 1.182 (0.788-1.771) |
| 5 Year | -0.156 | 0.239 | 0.428 | 0.513 | 0.855 (0.536-1.366) |
| Postgraduate | 0.044 | 0.249 | 0.032 | 0.859 | 1.045 (0.641-1.704) |
| Major (non-medicine) | 0.196 | 0.085 | 5.370 | 0.020 | 1.217 (1.031-1.436) |
| Marital status (unmarried) | 0.428 | 0.537 | 0.636 | 0.425 | 1.535 (0.535-4.399) |
| Only child or not (no) | 0.112 | 0.088 | 1.609 | 0.205 | 1.118 (0.941-1.329) |
| Academic performance (good ref) |  |  | 26.857 | < 0.001 |  |
| Poor | 0.852 | 0.182 | 21.929 | < 0.001 | 2.343 (1.641-3.347) |
| Moderate | 0.358 | 0.091 | 15.321 | < 0.001 | 1.430 (1.196-1.711) |
| Living expenses (more than needed ref) |  |  | 3.881 | 0.144 |  |
| Enough | 0.167 | 0.085 | 3.877 | 0.049 | 1.181 (1.001-1.395) |
| Inadequate | 0.129 | 0.142 | 0.820 | 0.365 | 1.137 (0.861-1.503) |
| Hours of sleep (less than 6 hours ref) |  |  | 30.520 | < 0.001 |  |
| 6~7 hours | -0.537 | 0.145 | 13.819 | < 0.001 | 0.584 (0.440-0.776) |
| 7~8 hours | -0.795 | 0.152 | 27.306 | < 0.001 | 0.451 (0.335-0.608) |
| More than 8 hours | -0.403 | 0.205 | 3.850 | 0.050 | 0.669 (0.447-1.000) |
| Stay up late or not (yes) | 0.060 | 0.153 | 0.155 | 0.694 | 1.062 (0.786-1.435) |
| Obsessive-compulsive symptoms<br>(Y-BOCS $\geq 16$) | 1.632 | 0.203 | 64.875 | < 0.001 | 5.115 (3.438-7.609) |

## Discussion

This study explored the prevalence of OCS in a representative sample of university students, addressing the gap of a lack of epidemiological studies in the post-pandemic era. In the present study, the probable prevalence of OCS was 6.5%. This finding was similar with studies done in Turkish 4.2% (Yoldascan et al., 2009), Saudi Arabia 5.1% (Sultan et al., 2021), India 8.5% (Jaisoorya et al., 2017), but lower than that reported in Iran 32.4% (Assareh et al., 2016) or America 35.7% (Huz et al., 2016). The possible discrepancy may be due to the different assessment criteria for obsessive-compulsive symptoms. The utilized criteria in Iranian research focused on subclinical symptoms, and the study conducted in America classified participants as having OCS based on the presence of at least one self-reported obsessive-compulsive symptom.

A meta-analytic review indicated that women are typically at greater risk of experiencing OCS than men (Fawcett et al., 2020). Consistent with previous studies, our results found that female students had a higher prevalence of OCS. Moreover, we found that the major (medical vs. non-medical) was related to obsessive-compulsive symptoms among university student samples. However, logistic regression revealed that medical students had a lower risk of OCS compared to non-medical students (OR, 0.680; 95%CI, 0.496-0.932), which is inconsistent with previous studies that found medical students were more prone to have obsessive-compulsive symptoms (Torres et al., 2016; Harries et al., 2017). The possible explanation may be that the non-medical students’ limited knowledge of OCD led to misinterpretation of obsessive-compulsive symptoms, potentially resulted in inflated self-reported symptom scores. As expected, a significant positive correlation was observed in this study between the family history of OCD and severity of OCS (total Y-BOCS score). A recent meta-analysis reported that OCD is a prevalent and highly familial disorder, which has a phenotypic heritability of around 50% among the relatives of children and adolescent probands (Blanco-Vieira et al., 2023). A review concluded that OCD has a significant hereditable component, with relatives of OCD cases being 4 times more likely to develop the disorder than the general population (Bloch and Pittenger, 2010).

Psychological strain can significantly predict suicidal ideation among Chinese and American college students (J. Zhang et al., 2017). In addition to the well-documented association between psychological strain and suicidal behaviors (Song et al., 2019; Tang et al., 2018; Wang et al., 2021), it is also reported that psychological strain was significantly related to depression (Zhang and Lv, 2014). These findings indicated that psychological strain is one of the significant contributing factors to the development of mental disorders. The severity of OCS was associated with coping strain in our study. More severe obsessive-compulsive symptoms were associated with higher level of coping strain scores. Severe and intense coping strain can cause mental disorders (Zhang et al., 2014), which may further exacerbate preexisting OCS. Notably, more severe obsessive-compulsive symptoms were associated with higher coping strain scores as well. Within non-clinical populations, subclinical OCS often received limited clinical attentions. The absence of professional guidance left individuals without the ability to control their symptoms, which is one of the reasons why the obsessive-compulsive symptoms worsen. The coping strain intensifies when limited self-control capacity that generates strong discomfort during the emergence of obsessions or compulsions clash with motivated attempts to suppress obsessive-compulsive symptoms. And these make the strong relationship between psychological strain and obsessive-compulsive symptoms more plausible.

## Implications for Practice

This study reported the prevalence of obsessive-compulsive symptoms (OCS) in Chinese university students and the association between psychological strain and obsessive-compulsive symptoms. The figure of 6.5% indicated that OCS is common in this population. The association between OCS and coping strain probably drives individuals toward extreme solution, such that even subclinical OCS may exacerbate psychological strain. Conversely, strong psychological strain may itself trigger OCS through inward frustration-aggression pathways. This vicious cycle suggests that the presence of OCS in university students may be a marker of experiencing psychological strain, which, could potentially escalate into serious mental disorders or suicide (Zhang et al., 2014). Universities and society should not overlook OCS, and mental health service providers should support university students in recognizing and managing OCS to help unlashing psychological strain for promoting overall mental health.

## Limitations

First, the self-reported data collection may involve information bias despite it being commonly used in the epidemiological study and multivariable regression model used. Second, psychological strain is predominantly investigated within the strain theory of suicide, with relatively scarce research exploring its association to other mental disorders. Consequently, our interpretations of the underlying mechanisms are likely to be less comprehensive. Third, most participants in this study were medical students, with fewer from other majors, which may limit the generalizability of the findings to university students in a specific major.

## Conclusions

Obsessive-compulsive symptoms (OCS) is a common mental condition among Chinese university students, with a prevalence of 6.5%. OCS is more frequent in university students reporting a family history of obsessive-compulsive disorder (OCD). A significant association exists between OCS and psychological strain, suggesting these constructs may mutually reinforce and maintain each other through a vicious cycle.

## Data statement

The datasets generated and/or analyzed during the current study are not publicly available due to institutional restrictions, but are available from the corresponding author upon reasonable request.

## Acknowledgments

The authors sincerely thank all study participants and research staff who have contributed to this work.

